# BodyMAE: A Surface-Area Aware Masked Autoencoder for Body Composition Estimation from 3D Body Scans

**DOI:** 10.64898/2026.06.04.26354925

**Authors:** Yijiang Zheng, Boyuan Feng, Ruting Cheng, Chuhui Qiu, Zhuoxin Long, Khashayar Vaziri, James Hahn

## Abstract

Accurate assessment of body composition is important to risk stratification and management of metabolic, musculoskeletal, and aging-related diseases, yet reference modalities such as Dual-energy X-ray absorptiometry (DXA) are costly and impractical for frequent monitoring. Commodity 3D body scans offer a low-cost, radiation-free alternative, but extracting meaningful and predictive shape features from scans remains challenging due to nonuniform point density, variable body size and cross-device differences. We introduce BodyMAE, a self-supervised, surface-area aware masked autoencoder for metric-scale 3D body scans. The pipeline integrates area-adjusted sampling, a long-range focused encoder, and a lightweight decoder regularized to promote locally uniform reconstructions. Trained and evaluated on 917 paired 3D body scans paired with clinical DXA reports, BodyMAE achieves strong accuracy on fat percentage (root-mean-square error (RMSE) 3.825 percentage points, R² 0.908), fat mass (RMSE 3.694 kg, R² 0.968), and lean mass (RMSE 3.608 kg, R² 0.901), with competitive performance on bone mineral content (RMSE 0.284 kg, R² 0.754).We also assess feature stability across pretrained baselines, finding higher retrieval accuracy for our representations (Top-1 90.131%). These results indicate that combining metric-aware sampling, long-range relational encoding, and local geometric regularization enables accurate body composition estimation from 3D body scans, as validated by comparisons to DXA-derived measurements.

## 1. Introduction

Metabolic-related diseases are strongly associated with body composition, especially the amount of body fat [1]. Higher total fat, and especially visceral adiposity, is linked to higher rates of hypertension and atherogenic dyslipidemia [2][3]. Nonalcoholic fatty liver disease (NAFLD) tracks with composition phenotypes, notably higher visceral fat and lower appendicular lean mass [4][5]. Moreover, declines in bone mass and bone quality elevate the risk of osteoporosis and consequent fragility fractures [6]. For sarcopenia, skeletal muscle mass, the muscle quantity, is a key diagnostic component, alongside muscle strength and physical performance [7]. In older adults, low muscle mass and density can increase the risk of falls, frailty, and mortality [8]. Therefore, evaluating body composition has direct clinical utility by refining risk stratification through quantifying fat mass, lean mass and bone health, which in turn guides prevention, pharmacotherapy, and treatment monitoring [9].

Developing affordable, accessible methods for accurate body composition estimation is an active research focus [10] [11] [12]. Traditionally, CT, MRI, and Dual-energy X-ray absorptiometry (DXA) are considered reference standards for regional and whole-body composition [13], yet CT and DXA involve ionizing radiation and all three are expensive and limited in availability, which constrains convenience and frequent monitoring [14]. These barriers motivate methods that work outside clinical settings. Optical 3D body scan addresses this gap by offering fast, low-cost, and radiation-free capture suitable for population-scale monitoring [15] [16]. Consumer-grade applications can deliver high-quality at-home scans using smartphone or tablet cameras, enabling detailed anthropometric measurements, such as waist and hip circumferences within minutes [17]. When paired with machine learning models that map shape to composition targets [11][12], these low-cost, repeatable scans enable reliable, high-frequency longitudinal tracking in both clinical and home settings.”

Unlike simple anthropometrics, extracting rich, predictive shape information from 3D body scans is challenging. Many approaches reduce the 3D surface to 1D anthropometric measures (e.g., calf or thigh circumference), discarding most geometric detail [18][19]. However, even circumference extraction from 3D body scans is nontrivial. Small slice offsets, axis misalignment, landmark ambiguity, and artifacts from limb contact or pose variation can all distort results. Fig. 1 shows typical contact cases where the arms touch the torso and the thighs touch each other, producing merged surfaces that distort cross-section contours and bias circumference estimates. Arm posture, including twisting or bending, further complicates accurate measurement. These challenges highlight the limitations of feature extraction based solely on predefined measurements or generic vision pipelines that ignore anatomical structure and metric scale, motivating approaches that learn directly from full 3D shape rather than reducing the surface to a few one-dimensional measures. Moreover, the 3D vision literature primarily focuses on classification and segmentation, leaving regression tasks such as body composition estimation underexplored [20]. Beyond circumference extraction, 3D body shape is typically represented as either a mesh or a point cloud. Mesh-based approaches often constrain scale and require extensive preprocessing for real-world scans. High-resolution meshes are memory-intensive, and many mesh methods assume watertight, manifold surfaces which require remeshing, hole filling, and topology cleanup [21][22]. In contrast, point cloud representations avoid topology constraints, scale naturally from thousands to millions of samples, and are easier to preprocess for raw body scans. Consequently, learning shape informative features directly from 3D body scan point clouds is a key approach to estimate body composition from these scans.

**Figure 1.**
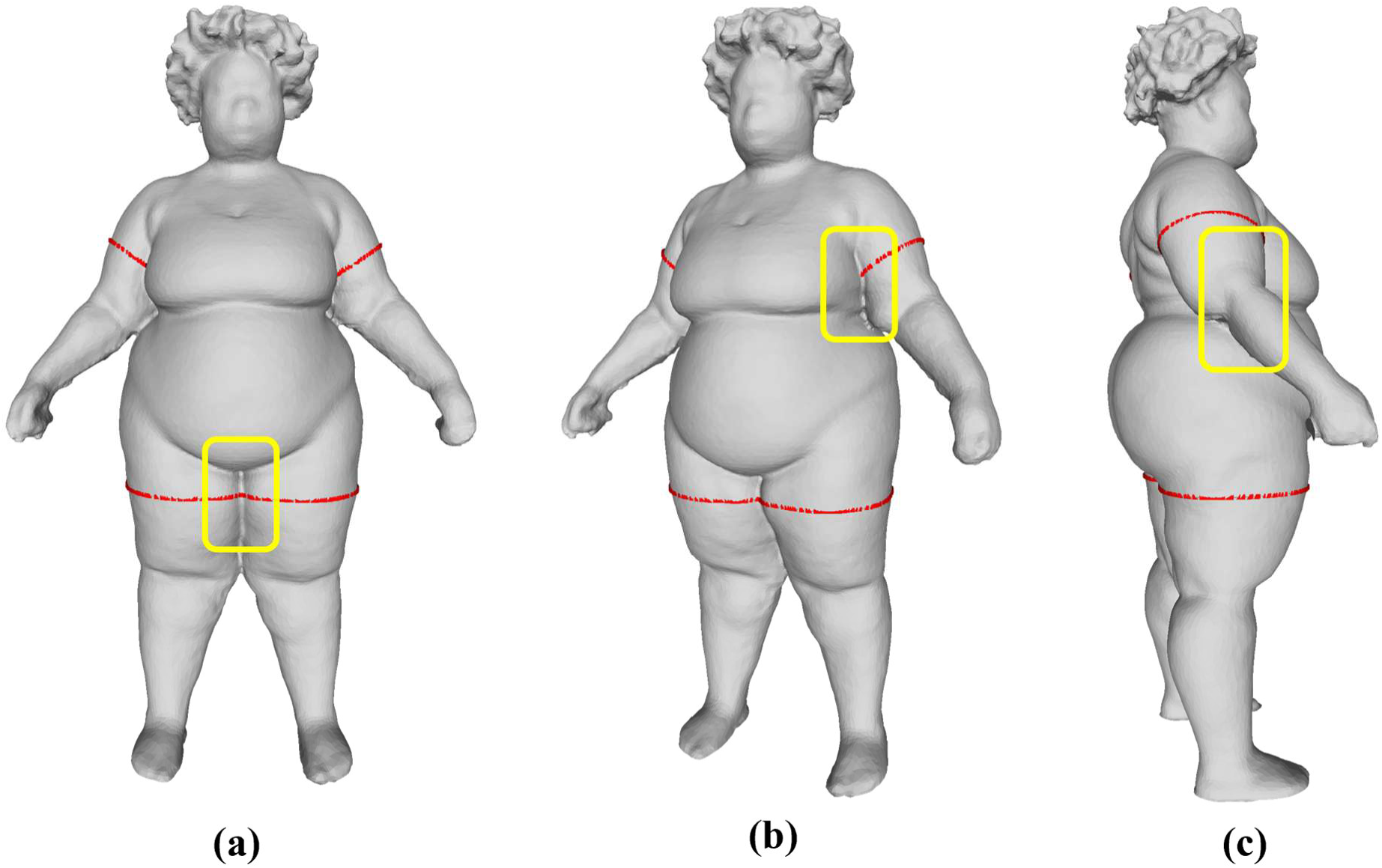
Sources of error in circumference extraction from 3D body scans due to limb contact and pose variation. Red dotted lines indicate intended measurement planes; yellow boxes highlight problematic regions. (a) Thighs touch and form a merged surface, making thigh girth ambiguous. (b) The upper arm contacts the torso, distorting both the arm and trunk contours. (c) A bent arm that is not aligned with the front arm shifts the cross section and changes the measured girth.

## 2. Related work

Several studies have applied 3D body scans for body composition estimation. Scan-derived anthropometric measures show strong agreement with whole body composition, particularly waist and hip circumference [10]. Methods that register scans to a human body template have used the resulting vertex correspondence to extract ordered features and apply principal component analysis (PCA) to these aligned shapes for predicting fat mass [23]. More recently, point cloud models with transformer architectures have reported improved correlations with whole and regional fat percentage [11]. In parallel, a generative line of work uses 3D body scans to synthesize pseudo-DXA images, enabling composition analysis when DXA is unavailable [24]. Despite these advances, labeled medical datasets remain small. Therefore, pretraining on large unlabeled 3D scans becomes crucial and consistently improves downstream body composition accuracy [12]. However, many prior works rely on low-resolution inputs or aggressively downsample the 3D shape to reduce computation. This approach risks discarding fine-grained geometric details that are critical for accurate composition estimation.

Point cloud deep learning has advanced rapidly in recent years. PointNet (PN) introduces permutation invariant set functions (e.g., max pooling) to handle unordered point sets, and PointNet++ (PN++) adds hierarchical neighborhood grouping and sampling to capture local geometry [25][26]. Sparse (Minkowski) CNN quantizes points into voxels and applies sparse 3D convolutions, yielding fast, memory-efficient models widely used in real-time autonomous driving perception [27]. DGCNN models a point cloud as a dynamic graph and applies edge convolutions, but rebuilding neighborhoods at each layer is memory and computation intensive, making it less suitable for dense human body scans [28]. Transformers have also been applied to 3D points. Point Transformer (PTv1) employs attention with relative positional encoding for local feature aggregation, while newer variants such as Point Transformer v3 (PTv3) scale attention to large point sets and more efficiently capture long-range dependencies by imposing an approximate spatial ordering [29][30].

Alongside model innovations, self-supervised learning is increasingly adopted in point cloud research for avoiding labor-intensive labels. Extensive studies show the later finetuning model which using self-supervised pretraining demonstrates superior performance than models training from scratch [31][32][33]. Point-BERT tokenizes local patches (via a learned codebook) and performs masked token prediction in a BERT-style objective [31]. Point-MAE partitions a cloud using farthest point sampling (FPS) for patch centers and K-Nearest Neighbors (KNN) for grouping, masks a high fraction of patches, and reconstructs the masked patches by predicting point coordinates normalized around each patch center [32]. Point-M2AE extends masked reconstruction with a U-Net like hierarchical encoder and decoder, but on dense human body scans, the large point counts make cross-scale neighborhood construction computationally and memory intensive [33]. Contrastive approaches create two augmented views of the same point cloud (e.g., rotations, scaling, jitter) as positives and push them together in embedding space while separating negatives [34].

Most point cloud pretraining targets object classification and segmentation on synthetic computer-aided design (CAD) shapes, which differ significantly from real human body scans used for body composition estimation [35][36]. Human anatomy is deformable, and scans can exhibit subtle soft-tissue displacement and sensor noise, whereas synthetic CAD models remain rigid, clean, and topologically simple. A common preprocessing step in point cloud work is global normalization (e.g., scaling each shape to a unit sphere or box), but this discards absolute metric scale, which matters for composition since larger bodies typically have greater component mass. Human body scans also have far higher point counts and nonuniform density across regions (e.g., trunk vs. limbs).

Differences in scanners and registration pipelines also introduce variability in resolution and geometric detail. As a result, common downsampling methods struggle in this setting. FPS promotes geometric uniformity but ignores physical surface area, assigning the same number of points to subjects of different sizes and thus producing inconsistent physical spacing between sampled points [26]. Voxel-based sampling depends on the original input density and requires careful choice of voxel size, which is difficult to tune so that point distributions remain comparable across subjects. In addition, body composition is a regression task that requires fine-grained, anatomically aware features at calibrated physical scale, and it benefits from modeling long-range relationships across distant body regions. Such context is valuable because composition patterns are shared across the body regions. Signals such as trunk–limb balance and bilateral symmetry (e.g., left and right thigh) cannot be recovered from a single local patch.

Given the challenges of learning from real 3D body scans, we introduce BodyMAE, a masked autoencoder tailored to metric-scale human scans and high density point sets. Our contributions are:

1. We introduce surface-area aware sampling that equalizes physical point spacing across subjects by scaling sample counts with estimated body surface area, stabilizing patch statistics and enabling a fixed-patch transformer while preserving comparable geometric detail.
2. We adapt Point Transformer v3 as the backbone to capture long-range spatial and fine-grained geometric relationships from 3D body scans using consistent physical aggregation, enriching point features while remaining memory efficient.
3. We incorporate a Repulsion Loss during masked reconstruction to discourage point clustering and promote locally uniform coverage, complementing Chamfer Distance, preserving high-frequency contours, and stabilizing pretraining for sharper patch features.
4. We demonstrate improved prediction of body composition outcomes compared with other baselines, achieving lower RMSE and higher R² across total body fat percentage, regional fat and lean mass.

## 3. Method

Most optical body scanners produce surface meshes, and point clouds are routinely derived from these representations [26]. The point sets enable direct computation of point normal, surface area, regional circumferences, and volume (when the mesh is watertight), which in turn supports normalization, quality control, and downstream analyses.

### Area-adjusted FPS

FPS preserves the native coordinates and yields near-uniform, near-isotropic coverage (Fig. 2). In contrast, grid random sampling (GRS) first voxelizes the cloud and then selects one original point per occupied voxel, introducing intra-cell variance. Grid mean sampling (GMS) replaces all points within each voxel by the voxel centroid, effectively quantizing coordinates onto a regular lattice. However, GMS creates aliasing artifacts and displaces boundary points from their true locations. Consequently, FPS achieves more uniform within-scan spacing than grid-based samplings (see Experiment 1). Nonetheless, fixing the number of FPS points per scan produces uneven physical point density across subjects: larger bodies are under-sampled while smaller bodies are over-sampled. To maintain an approximately constant density and make masked-autoencoder reconstructions more comparable across subjects, we use surface area adjusted FPS. Because our scans are optical surface captures, density should be scaled with surface area, not volume. We set the target point count *N* by ideal hex-packing on the surface [37]. A hex cell with mean inter-sample spacing ℎ has area 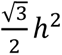, thus for a scan with surface area *S*:

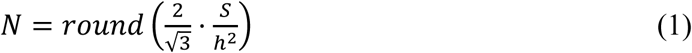

In practice we compute *S* as the sum of triangle areas after removing duplicate and degenerate faces. By adjusting the target spacing ℎ at each downsampling step, we set *N* using the same formula to enforce subject-invariant point density, producing comparable masked patches across body sizes.

**Figure 2.**
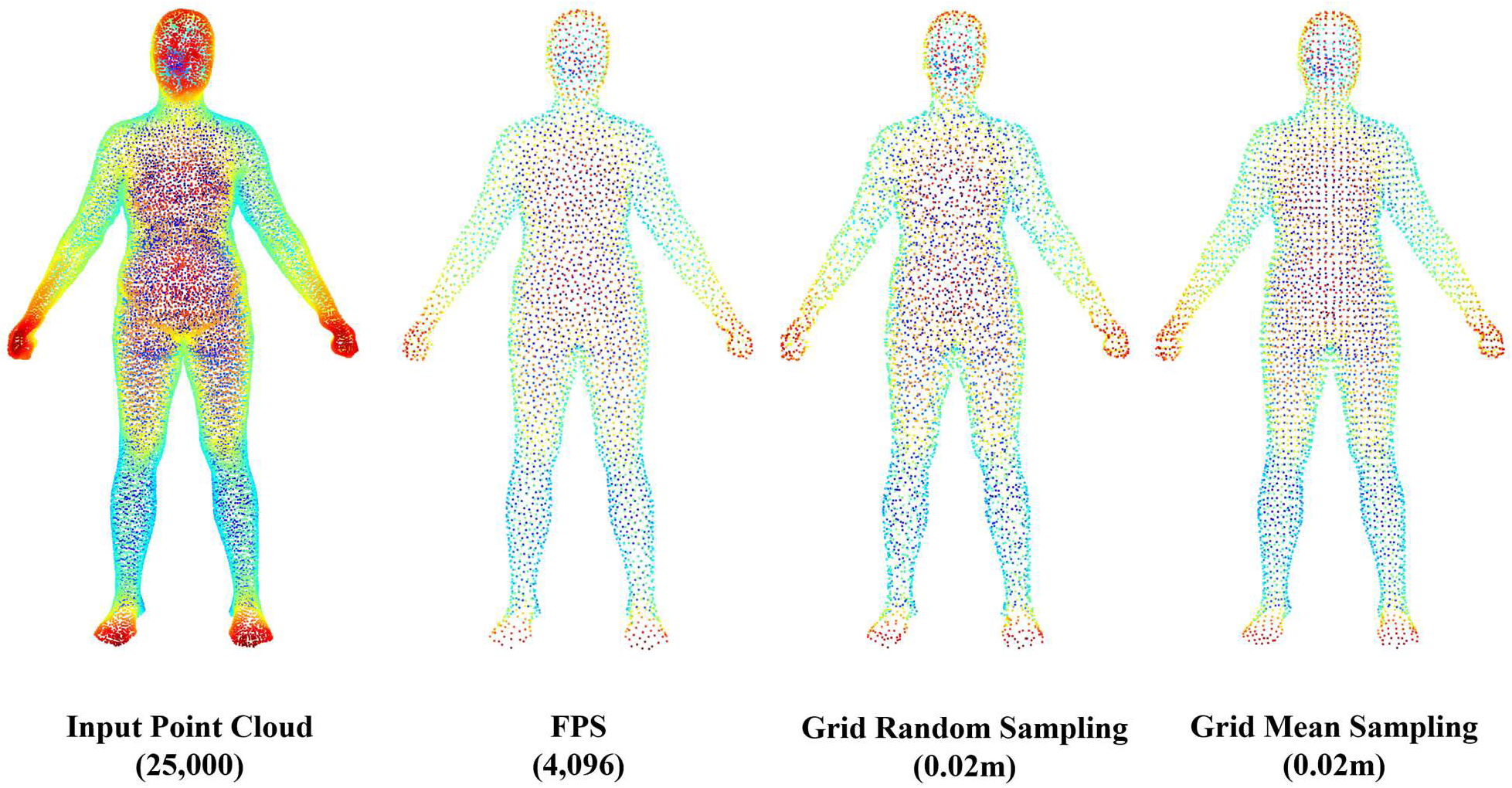
Comparison of downsampling strategies on the same scan. Left: input point cloud (25,000 points). Center-left: FPS (4,096 points) gives near-isotropic, uniform coverage. Center-right: GRS at 0.02 m (one point per occupied cell) yields uneven intra-cell spacing and axis-aligned artifacts. Right: GMS at 0.02 m (cell centroids) imposes a lattice but shifts boundary points, altering the native distribution. FPS best preserves the original local geometry and uniform density for subsequent MAE patching. Voxel size 0.02 m is set to yield ∼4k points, matching FPS.

#### BodyMAE

As presented in Fig. 3, we adopt PTv3 as a lightweight, point-centric encoder. Starting from the raw ∼25k points, the block embeds the coordinates and at each layer divides the cloud into large, non-local patches (the colored regions in Fig. 3 “Divide”). Points with the same color belong to one patch and attend to one another, so each point absorbs long-range context, including across bilateral structures such as the left and right legs. At each layer we also downsample the points to form the inputs for the MAE. We simply apply the previously introduced area-adjusted FPS with a metric radius Ball Query so patch density is physically consistent across subjects. This keeps the original coordinates intact while delivering uniform, comparable patches for the subsequent MAE stage.

**Figure 3.**
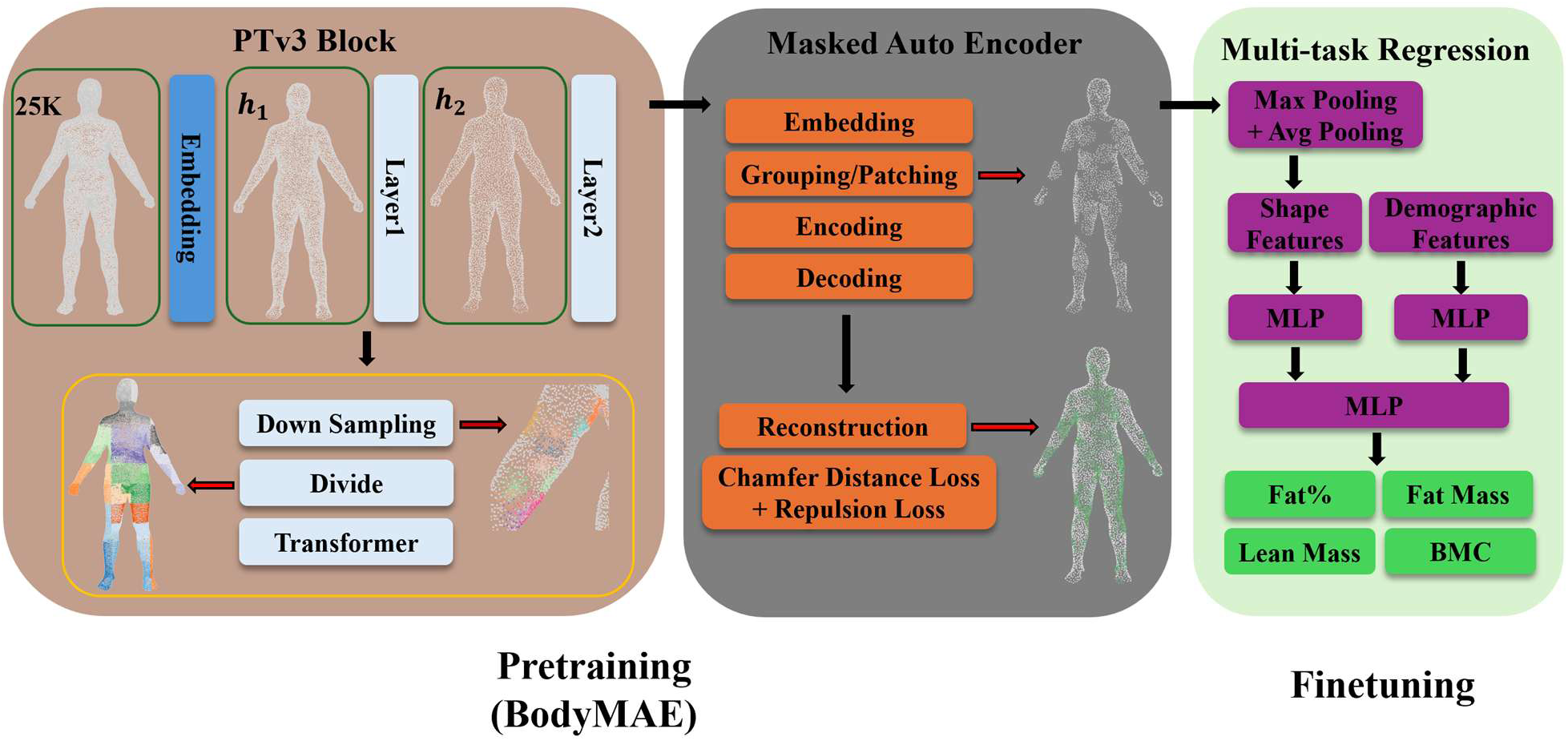
Overview of the proposed pipeline. Pretraining (BodyMAE): an input point cloud (25k) is processed by a PTv3 block to obtain hierarchical features at target spacings ℎ_l_ and ℎ_2_. Points are downsampled and divided into patches, which are embedded, encoded, and decoded in a masked autoencoder. Reconstruction uses symmetric Chamfer Distance with an added repulsion term to discourage local clustering. PTv3 block captures long-range context and MAE reconstructs masked patches. Finetuning: patch embeddings are aggregated by max and mean pooling to a subject level shape descriptor, fused with demographic features through separate MLP branches, and passed to a shared MLP that predicts four DXA targets: body fat percentage, fat mass, lean mass, and bone mineral content.

From the PTv3_block output, we partition the point set into 128 groups and around each center, gathering 64 neighbors using KNN. At this stage, every group has comparable physical support resulting from the previous area-adjusted FPS. The group points are presented as offsets relative to their center (as the same with MAE) and apply a fixed masking ratio of 0.7 over the 128 groups: visible groups are encoded, while masked groups are reconstructed by a lightweight decoder that predicts the 64-point patch back in the local frame. Training uses L2 Chamfer Distance Loss on coordinates (computed in the local frame with the usual scale handling), plus a light Repulsion Loss term to prevent intra-group clumping. This setup preserves original coordinates, enforces uniform group density, and makes the reconstruction objective consistent across subjects.

### Repulsion Loss

We apply a repulsion term at the decoder’s last reconstruction stage to discourage local clustering in the predicted points while preserving the global fit enforced by Chamfer Distance Loss [38][39].

For a single reconstructed patch, let 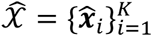 be the predicted points and define the distance between points ***x̂****_i_* and ***x̂****_j_* as 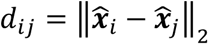. We set the penalty boundary distance as *r* = *α* ⋅ ℎ_2_, the ℎ_2_ is the target spacing distance from the final downsampling level. We set *α* as 0.5, so only pairs closer than half of the mean space distance are penalized. To focus the penalty on true local contacts and keep the normalization stable, we sum over the *k_top_* nearest neighbors of each point within the patch, rather than over all pairs. This yields stronger, more stable gradients (undiluted by distance from non-violating pairs) and reduces computation from *O*(*K*^2^) to *O*(*Kk_top_*). Therefore, the Repulsion Loss can be written as:

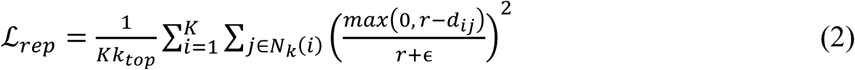

Where *∊* > 0 is a small stabilizer and *N_k_*(*i*) is the set of the *k_top_* nearest neighbors of point *i* inside the patch. The final loss function is the combination of the ℒ*_chamf_* and ℒ*_rep_*. In practice, the ℒ*_rep_* behaves as a localized regularizer that does not dominate ℒ*_chamfer_*, yielding stable optimization without tuning an extra weight.

### Body Composition Model

After pretraining, the BodyMAE encoder produces 128 patches shape features for each scan. Let *F* ∈ *R^P^*^×*C*^ denote the matrix of patch features (*P* patches, *C* channels). We form a subject-level shape descriptor *F_s_* via two pooling as:

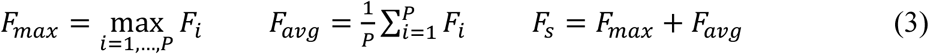

The max pooling features and mean pooling patch features are added to form the whole body shape descriptor *F_s_* [33]. Demographic features *F_d_* includes age, gender, ethnicity, height, weight, and BMI. We include BMI explicitly, even though it can be derived from height and weight, because providing it directly supplies a strong, stable signal. All demographic variables are standardized during the preprocessing. As the finetuning part demonstrated in Fig. 3, the shape descriptor *F_s_* and the demographics *F_d_* are each passed through two separate MLPs, and the results are concatenated into final feature *Z* before the regression head:

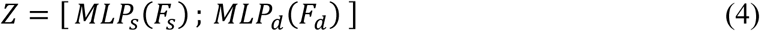

A shared MLP maps *Z* to four regression targets: body fat percentage (fat%), fat mass, lean mass, and bone mineral content (BMC). The head consists of fully connected layers with nonlinearity and dropout. We train the model by minimizing the mean squared error (MSE) averaged across tasks on standardized targets, and we do not apply task weights since standardization equalizes scale:

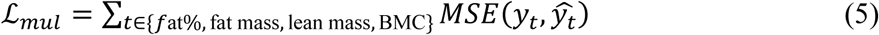

The four targets are physiologically coupled and driven by overlapping 3D shape information, so a shared encoder is statistically well-motivated. In our experiments, separate single-task models trained over multiple runs achieved accuracy similar to the multi-task model. Given these comparable results, we favor the multi-task setup for practicality: one model with shared hyperparameters and consistent cross-task representations, lower compute cost, and simpler deployment, while preserving the inductive bias of shared structure.

## 4. Dataset and experiment settings

All body composition datasets in this study are collected under the standard optical body scan protocol: standing A-pose with tight-fitting clothing. This can help reduce self-occlusion and clothing artifacts and ensure comparability across cohorts. We choose pretraining datasets that match this posture and modality, so the learned representation is aligned with the downstream body composition cohorts. These pretraining sets are larger and more diverse in body shape and provide consistent shape alignment, which reduces pose-related domain shift and improves transfer to the body composition datasets.

### Pretraining dataset

We include 3 dataset sources as our pretraining datasets:

- Semantic-parameter (SP) dataset [40]. Raw 3D body scans are non-rigidly registered to a standardized human template using semantic parameters, producing topology-consistent, spatially aligned meshes across subjects. Each mesh contains 12,500 vertices and 25,000 faces. The dataset comprises 3,048 scans.
- Caesar Dataset (CA) [41]. We use the publicly available processed CAESAR meshes, and it contains only the U.S. cohort (the European subset is not included). Each subject mesh has higher resolution (around 50,000 vertices) with consistent topology and alignment. The processed set includes 2,358 scans, and all results reported for CAESAR are therefore based on this U.S.-only subset.
- Obstetrics (OB) dataset [42]. Scans were acquired with a standard Fit3D scanner, which outputs meshes directly. As an optical, non-ionizing modality, Fit3D is suitable for use during pregnancy, unlike CT or DXA, which are generally not recommended. All scans were collected between 18–38 weeks’ gestation. Each mesh contains approximately 25,000 vertices, and the dataset includes 711 scans.

The pretraining corpus comprises 6,117 scans, combining consistent topology and alignment with broad shape diversity to support robust representation learning.

### Body Composition datasets

Each subject undergoes optical 3D surface scanning and DXA, yielding paired shape–composition data. The cohort spans a wide range of body types, including normal BMI adults, individuals with severe obesity evaluated pre-bariatric surgery, and older adults (≥65 y) in a sarcopenia cohort. All DXA examinations were performed on GE Lunar systems. Each session produced a composition report with three components: fat mass, lean mass and BMC, along with the derived body fat percentage, a key metric for routine monitoring. These measurements serve as the ground truth for training and evaluating our models.

DXA report examples are shown in Fig. 4. Notably, when the DXA table cannot accommodate individuals with obesity, clinical protocols recommend scanning only the right side and mirroring to estimate the left side’s composition [43]. This practice readdresses the importance of long-range contextual information when estimating body composition. We evaluate three cohorts:

- Normal BMI dataset (NB) [44]: 489 scans acquired with a high-resolution optical scanner (around 100,000 vertices per mesh).
- Bariatric dataset (BA) [45]: 252 scans from participants with BMI greater than 35 who were scanned before bariatric surgery. Meshes contain about 25,000 vertices from a standard Fit3D scanner.
- Sarcopenia dataset (SA): 176 scans focusing on older adults with age at least 65, also collected on a standard Fit3D scanner with meshes of about 25,000 vertices.

**Figure 4.**
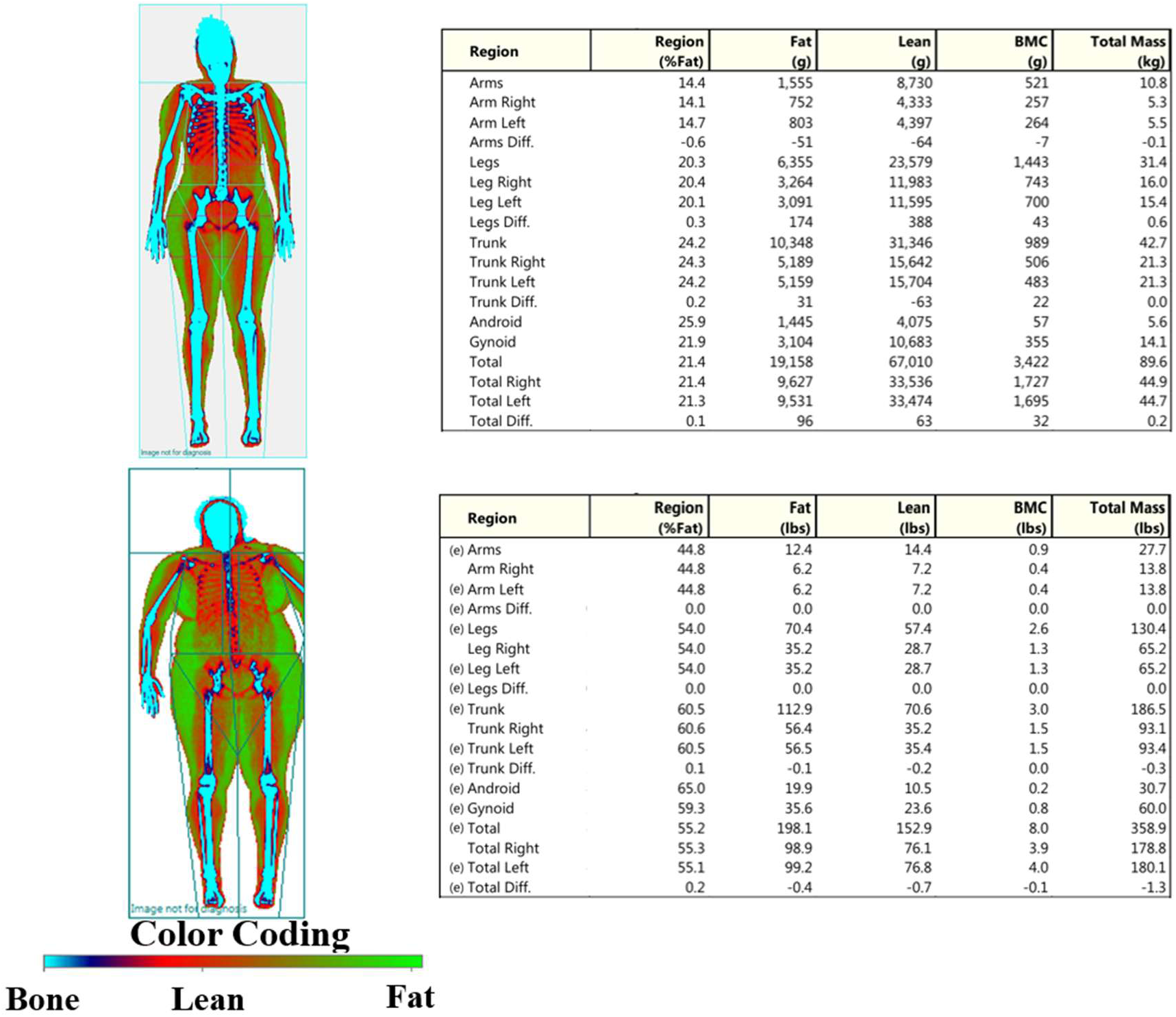
DXA body composition reports. Top: representative subject with normal BMI. Bottom: obese subject where, due to DXA table size limits, only the right side is scanned, and left-side values were estimated by symmetry (denoted by “(e)” preceding the region). The report includes fat percentage, fat mass, lean mass, and bone mineral content.

In total, the body composition dataset contains 917 that combine 3D body scans with DXA measurements across NB, BA, and SA datasets. All numeric variables are unified across cohorts: weight, fat mass, lean mass, and BMC are reported in kilograms; height in meters. A detailed summary is reported in the Supplementary Material.

### Experiment Settings

For self-supervised pretraining we hold out 20% of scans for validation. For the downstream task, we perform five-fold cross-validation stratified by cohort (NB, BA, SA) to assess generalization and limit overfitting.

All pretraining and downstream 3D body scans are normalized to meter scale. Each scan is represented as a set of 25,000 points. If a mesh has at least 25,000 vertices, we uniformly subsample 25,000 vertices without replacement. If it has fewer vertices, we use all available vertices and then sample the remaining points uniformly on triangle faces with probabilities proportional to face area until reaching 25,000. Normal for vertex-sampled points come from the mesh vertex normal, and normal for face-sampled points are obtained by barycentric interpolation of the incident vertex normal. Surface area is computed during preprocessing as the sum of triangle face areas. Because subjects are scanned in a consistent standing A-pose, we apply only light geometric augmentation: small isotropic jitter, small translations, and a yaw rotation about the y-axis within ±15 degrees. All augmentations preserve metric scale, and normal features are re-normalized to unit length afterward.

The PTv3_block divides points into patches of size 512 at each layer. Hierarchical downsampling spacings ℎ_l_ and ℎ_2_ are 0.0141 and 0.02. For MAE, we form 128 groups with each group containing 64 points. For downstream evaluation, we report RMSE and R² for all regression targets.

Models are trained for 300 epochs with AdamW (learning rate 0.001), a cosine learning rate schedule with a 10 epochs warmup, and batch size 32. We evaluate the validation set every 10 epochs and keep the checkpoint with the best validation performance. This protocol follows common practice in prior work [30][31]. All experiments are implemented in Pytorch and run on four NVIDIA RTX 6000 Ada GPUs.

## 5. Results & Discussion

### Sampling Methods distribution and density

In this experiment, for the FPS baseline we fix the point count *N_p_* to 4096 for every scan. Using Eq.(1), a single reference spacing ℎ*_ref_* is obtained from the dataset mean surface area *S̅*:

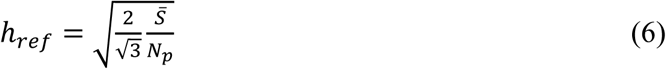

Then we apply the ℎ*_ref_* to parameterize area-adjusted FPS on all scans. To compare with the grid-based sampling while matching point density per surface area, we set the grid cell size *c* to have the same area per point as hex packing with ℎ*_ref_*:

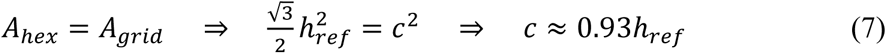

This ensures the grid produces a comparable density, allowing a fair comparison of distribution quality. To evaluate sampling density within and across subjects (scale-aware), we compute the per-scan mean nearest-neighbor distance *μ_s_* and report its across-subject standard deviation *σ_µ_*. We also report within-scan uniformity via the mean of coefficient of variation 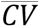:

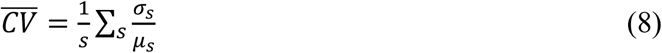

where the *σ_s_* is the within-scan standard deviation of nearest-neighbor distances. Lower *σ_µ_* indicates better cross-subject density consistency and lower 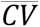 indicates more uniform spacing within scans. We also report coverage of each sampled set relative to the 25k input cloud from the same scan. Coverage is measured with the Chamfer Distance. A lower value indicates that the sampled points more completely cover the original surface geometry (fewer holes and smaller boundary shifts).

From each dataset (SP, CA, OB, NB, BA, SA), we randomly sample 100 scans, yielding a total of 600 scans for testing. Fig. 5 compares the sampling methods in three metrics. Area-adjusted FPS achieves the lowest cross-subject *σ_µ_* because the sample count scales with surface area, keeping physical spacing stable across different body sizes, whereas fixed point count FPS varies with subject size and grid methods cannot match density across subjects due to geometry dependent cell occupancy. FPS and area-adjusted FPS also yield lower 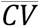 than grid sampling. GRS leaves offsets within occupied cells and GMS imposes a lattice pattern, both of which create uneven nearest-neighbor distance. For coverage, GMS shifts boundary points toward cell centroids and increases Chamfer Distance to the original points, while other methods achieve similar lower values. Taken together, area-adjusted FPS provides the best cross-subject consistency, strong within-scan uniformity, and good coverage. Accordingly, we adopt it as our downsampling strategy to produce more uniform MAE patches and improve reconstruction.

**Figure 5.**
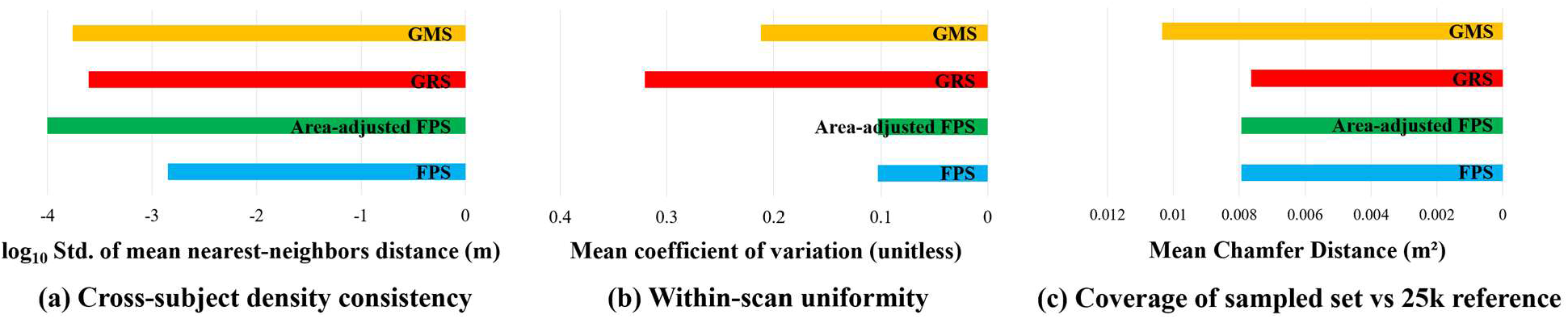
Comparison of sampling strategies on body scans. (a) Cross-subject density consistency, shown as the log-scaled (compress the dynamic range for clearer comparison) standard deviation of mean nearest-neighbor distances. (b) Within-scan uniformity, measured by the mean coefficient of variation. (c) Coverage relative to the 25k-point input point cloud, quantified by mean Chamfer Distance. Lower values indicate better performance for all metrics. GRS: grid random sampling; GMS: grid mean sampling.

### Body Feature Similarity

Stability is critical for reliable body composition prediction. Features should remain consistent across repeated scans of the same subject, even when demographic metadata is unavailable. Our downstream task set includes subjects with multiple back-to-back scans (1,226 scans forming paired comparisons), enabling a direct assessment of feature consistency. We compute cosine similarity between feature embeddings of repeated scans and quantify stability using Top-1 and Top-5 accuracy metric. Comparative results between our pretrained model and popular alternatives are summarized in Table 1.

**Table 1.**
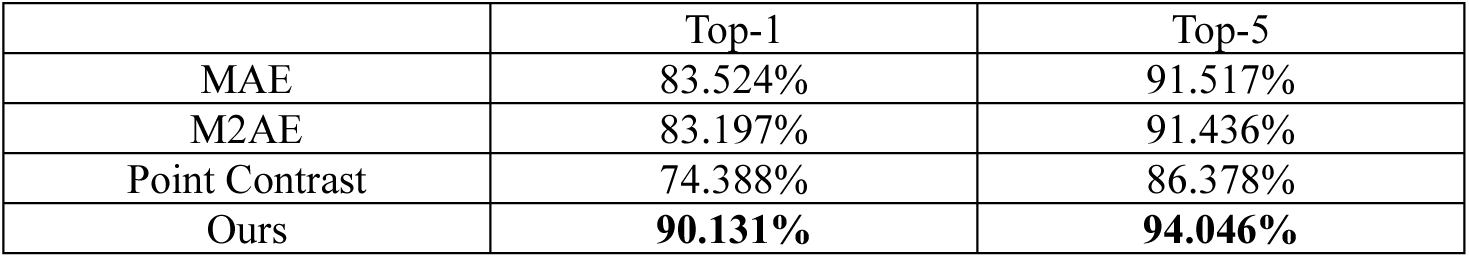
Pretrained Model Comparison on Body Feature Similarity (Top-1/Top-5 Accuracy).

The results highlight clear differences in feature stability across pretraining strategies. Both MAE and M2AE achieve strong performance, with Top-1 accuracies around 83% and Top-5 near 91%, indicating that masked autoencoding approaches capture relatively consistent intra-subject representations. Point Contrast performs notably worse, with only 74.388% Top-1 and 86.378% Top-5, suggesting that contrastive learning alone is less effective at preserving stability across repeated scans. Masked autoencoding methods encourage the model to learn global structure and spatial continuity of human body scans, leading to more consistent features. In contrast, contrastive learning treats each scan as a distinct instance, which can exaggerate minor variations and reduce the representation stability. Our method achieves the highest stability, reaching 90.131% Top-1 and 94.046% Top-5, outperforming all baselines by a significant margin. This demonstrates that our approach strengthens feature consistency across repeated scans compared to existing pretraining frameworks, providing a more robust foundation for body composition prediction.

### Body Composition Evaluation

We compare our method against a range of baselines. The Demographic Features (DF) model uses only basic subject metadata and does not incorporate any 3D shape information. The Level-Circumference (LC) model represents body shape by extracting multiple cross-sectional girths (“level” slices) from the limbs and torso [16][46]. Models highlighted in red indicate using self-supervised pretraining.

In Table 2, the body composition comparison highlights several key findings. Demographic features provide a reasonable baseline (e.g., R² = 0.903 for fat mass), indicating that basic human characteristics capture part of the variance in body composition. However, DF performs relatively poorly for the other components, emphasizing that demographics alone are insufficient to represent body shape or accurately estimate composition. LC models yield modest improvements over DF (e.g., fat% RMSE = 4.587 percentage points vs. 4.985), showing that regional circumferences introduce useful geometric information but remain too coarse to capture deeper composition differences.

**Table 2.** Models Performance on Body Composition Outcomes: Fat%, Fat Mass, Lean Mass, and BMC (RMSE/R²).

|  | Fat% |  | Fat Mass |  | Lean Mass |  | BMC |  |
| --- | --- | --- | --- | --- | --- | --- | --- | --- |
| | RMSE | $R^2$ | RMSE | $R^2$ | RMSE | $R^2$ | RMSE | $R^2$ |
| DF | 4.985 | 0.847 | 6.447 | 0.903 | 5.076 | 0.803 | 0.340 | 0.655 |
| LC [16] | 4.587 | 0.872 | 5.999 | 0.916 | 4.313 | 0.858 | 0.319 | 0.690 |
| PN [25] | 4.775 | 0.862 | 5.108 | 0.939 | 4.135 | 0.869 | 0.319 | 0.691 |
| PN++ [26] | 4.522 | 0.876 | 5.931 | 0.918 | 4.413 | 0.851 | 0.308 | 0.712 |
| Mink [27] | 4.715 | 0.865 | 6.015 | 0.915 | 4.189 | 0.866 | 0.290 | 0.743 |
| PTv1 [29] | 4.452 | 0.880 | 5.969 | 0.917 | 4.148 | 0.869 | 0.309 | 0.709 |
| PTv2 [47] | 4.460 | 0.879 | 5.655 | 0.925 | 3.823 | 0.888 | 0.294 | 0.738 |
| PTv3 [30] | 4.513 | 0.876 | 5.818 | 0.921 | 3.923 | 0.882 | 0.289 | 0.746 |
| **MAE [32]** | 4.748 | 0.863 | 4.287 | 0.957 | 4.304 | 0.858 | **0.277** | **0.766** |
| **M2AE [33]** | 4.316 | 0.887 | 4.408 | 0.955 | 3.810 | 0.889 | 0.279 | 0.763 |
| **Point Contrast [34]** | 5.205 | 0.836 | 7.102 | 0.882 | 4.769 | 0.826 | 0.322 | 0.684 |
| **Ours** | **3.825** | **0.908** | **3.694** | **0.968** | **3.608** | **0.901** | 0.284 | 0.754 |

Incorporating 3D body scans with point-based methods markedly enhances prediction accuracy across all traits, with PN++ achieving R² = 0.876 for fat% and PN excelling in fat mass. This demonstrates that geometric information from scans provides complementary value beyond demographics, particularly for fat-related measures. Minkowski networks further improve BMC estimation (R² = 0.743), highlighting the utility of voxelized sparse convolution for capturing skeletal structure. Transformer-based models (PTv1–3) achieve stronger performance in lean mass prediction (e.g., PTv3: R² = 0.882), indicating that long-range relational modeling is effective for distributed muscular patterns. Nevertheless, these approaches still fall short of mask-based self-supervised methods.

Pretraining generally improves downstream performance, particularly in the medical domain [48][49]. In our experiments, masked autoencoders (MAE, M2AE) substantially outperform non-pretrained architectures, especially in fat mass prediction, with MAE also achieving the best result in BMC. This confirms that reconstruction-based self-supervision learns robust structural features that transfer effectively to body composition tasks. In contrast, Point Contrast performs weakest (R² = 0.826 for lean mass, 0.882 for fat mass), highlighting its instability for body scans where small pose or scan variations undermine contrastive objectives. Our method achieves the best results in fat%, fat mass, and lean mass, with fat% RMSE reduced to 3.825 percentage points (R² = 0.908), fat mass to 3.694 kg (R² = 0.968), and lean mass to 3.608 kg (R² = 0.901). These correspond to ∼23.2% and ∼12.8% relative error reduction in fat% compared with the DF baseline and the strongest pretrained comparison (M2AE), respectively. The improvements are especially pronounced in fat-related outcomes, which are most clinically relevant for obesity and metabolic disease. Although our model does not surpass MAE in BMC prediction (RMSE = 0.284 kg vs. 0.277 kg), it remains competitive. Notably, all models yield relatively lower R² for BMC, suggesting that fat and lean mass are more closely tied to skeletal variation and demographics, whereas BMC is less directly reflected in surface geometry.

Overall, these findings demonstrate that demographic features alone offer only a coarse approximation of body composition, while 3D body scans, especially when paired with advanced pretraining, capture the subtle geometric details needed for accurate fat and lean mass estimation. By combining area-adjusted sampling, PTv3-based long-range encoding, and geometric regularization, our approach achieves state-of-the-art accuracy and stability.

### Ablation Study

#### Impact of PTv3_block and Repulsion Loss

To assess the effectiveness of our proposed modules, we conduct an ablation study examining the impact of PTv3_block and Repulsion Loss on body composition prediction.

From Table 3, adding either module improves performance, and using both yields the best overall body composition prediction except for BMC, where adding only Repulsion Loss is strongest. Relative to the baseline, PTv3_block most benefits fat mass (RMSE ↓17.7%), with smaller gains elsewhere. Repulsion Loss more strongly benefits fat% and lean mass (RMSE ↓6.0% and ↓10.5%) and also modestly improves BMC. Combining both modules delivers the best scores for fat%, fat mass, and lean mass. Thus, both modules are necessary for a balanced, high accuracy system. PTv3_block supplies long-range geometric context that stabilizes mass estimates, while Repulsion Loss enforces local geometric regularity that sharpens fat and lean predictions, together they provide complementary gains that generalize across targets, with the minor caveat that BMC may favor Repulsion Loss alone.

**Table 3.** Effect of PTv3_block and Repulsion Loss (with vs. without) on Body Composition Prediction: Fat%, Fat Mass, Lean Mass, and BMC.

| PTv3_block | RL | Fat% |  | Fat Mass |  | Lean Mass |  | BMC |  |
| --- | --- | --- | --- | --- | --- | --- | --- | --- | --- |
| | | RMSE | $R^2$ | RMSE | $R^2$ | RMSE | $R^2$ | RMSE | $R^2$ |
|  |  | 4.407 | 0.878 | 4.781 | 0.942 | 4.262 | 0.861 | 0.289 | 0.746 |
| ✓ |  | 4.352 | 0.885 | 3.936 | 0.960 | 4.096 | 0.872 | 0.288 | 0.747 |
|  | ✓ | 4.144 | 0.896 | 4.470 | 0.949 | 3.814 | 0.889 | 0.282 | 0.759 |
| ✓ | ✓ | 3.825 | 0.908 | 3.694 | 0.968 | 3.608 | 0.901 | 0.284 | 0.754 |

#### Effect of Patch Configuration on Accuracy and Latency

We also examine how group count and neighbor count affect average R² and inference latency across different settings (Fig. 6).

**Figure 6.**
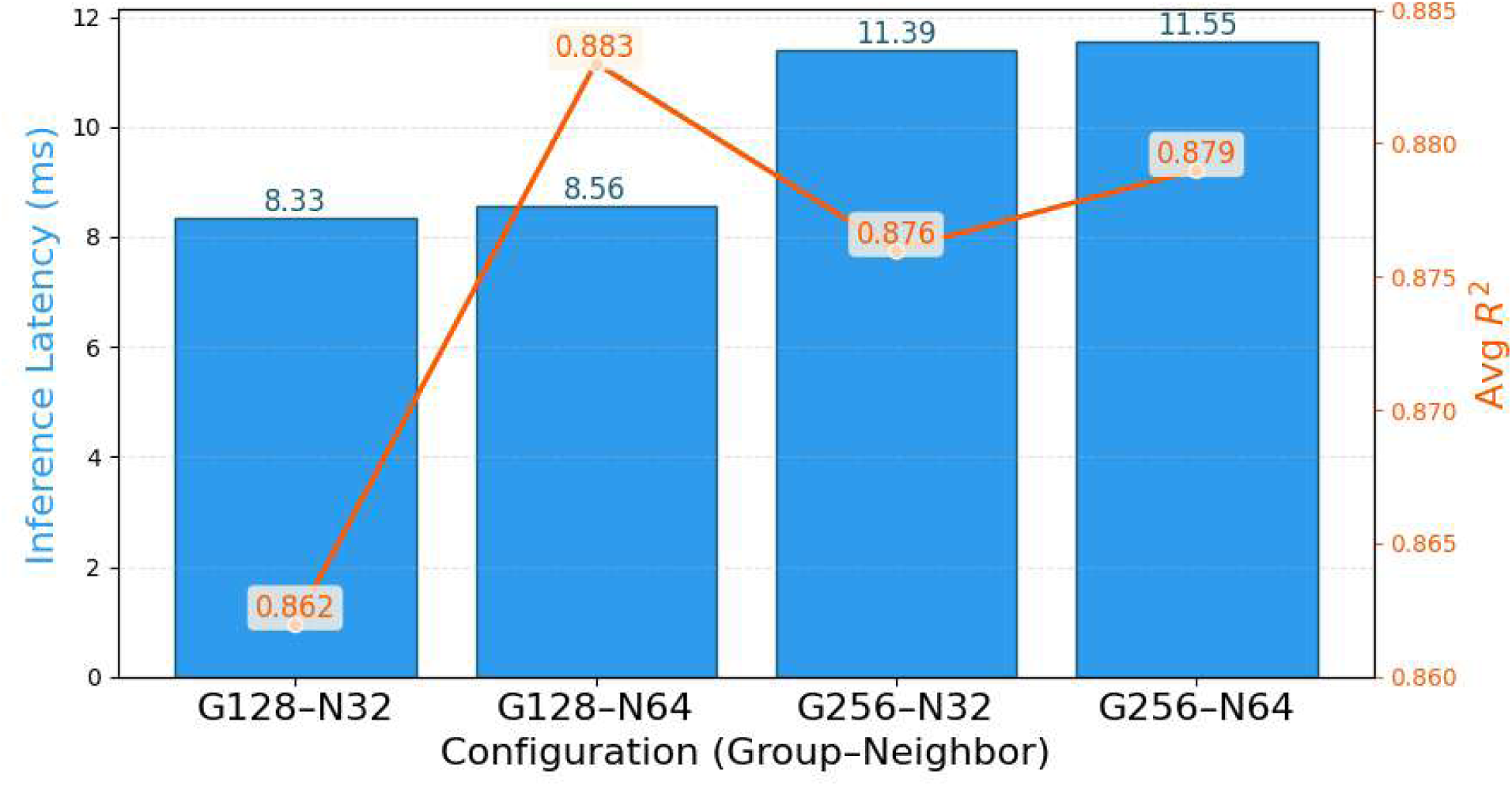
Effect of patch configuration on inference latency and accuracy. Bars show inference latency (ms) measured at batch size=1, while the line plots the average R² across Fat%, Fat Mass, Lean Mass, and BMC. Here, G denotes the group (patch) count and N the neighbor count.

The patching ablation shows that inference time is driven mainly by the number of groups (patch count), while accuracy is most sensitive to the number of neighbors. Configurations with 128 groups are consistently faster than those with 256 groups (around 8.3-8.6 ms vs. around 11.4-11.6 ms) and increasing neighbors from 32 to 64 adds only a small latency bump within a fixed group count. In contrast, average R² improves when increasing neighbors, e.g. 0.862→0.883, but moving from 128 to 256 groups does not guarantee to yield accuracy gains despite the higher cost. Overall, G128–N64 offers the best accuracy and efficiency trade-off, achieving the highest R² (0.883) with only a modest increase in latency over the setting G128–N32.

## 6. Conclusion

We presented BodyMAE, a surface-area aware masked autoencoder tailored to metric scale 3D body scans for body composition estimation. Our experiments use several datasets of 3D body scans paired with DXA reports, providing ground truth for fat%, fat mass, lean mass, and BMC and enabling direct evaluation. By coupling area-adjusted FPS with a PTv3-based encoder and a lightweight decoder regularized by Repulsion Loss, our method learns stable, scale-aware shape features and achieves superior accuracy across fat%, fat mass, and lean mass, while remaining competitive on BMC. The sampling strategy maintains uniform physical point density across different sized bodies, preserves consistent patch scale and reduces lattice artifacts that can distort local neighborhoods. This yields more stable nearest-neighbor statistics, improves MAE patch reconstruction by feeding comparably dense patches, and stabilizes training by avoiding over or under sampling of different body regions. Additionally, the proposed model has better feature stability (Top-1/Top-5 retrieval accuracy) compared to other pretraining models. For downstream prediction, models with mask-based pretraining consistently outperform non-pretrained baselines across all body composition endpoints. Together, these results indicate that metric-aware sampling, long-range relational encoding, and local geometric regularization are complementary for extracting clinically meaningful information from 3D body shape.

Our work still has several limitations. First, the paired 3D–DXA cohort, while demographically varied, comes from a limited number of acquisition sites, which risks site-specific biases (operator protocols, background geometry) and limits external validity. Second, scanner heterogeneity is only partially covered. Differences in sensor type (structured light vs. LiDAR), calibration, depth noise, mesh denoising, and coordinates conventions can alter point density and local surface statistics, potentially degrading performance when deployed on unseen devices or software versions. In addition, despite standardized posture guidance and area-aware sampling, residual domain shifts persist from clothing and hair, self-occlusions, stance variability, and registration pipeline choices (template fitting, hole filling, smoothing, normal estimation), each of which can subtly distort geometry and downstream features. Collectively, these factors mean the learned representations may be limited to sites, scanners and processing pipeline, and performance on newer scanners, processing stacks, or underrepresented populations may not match the reported results without additional adaptation or calibration.

Future work should include multi-center, cross-device validation with larger cohorts and standardized protocols for generalization purposes, especially in at-home capture settings relevant to telehealth. It will also be important to incorporate richer reference standards (e.g., MRI-derived visceral adiposity, CT-based bone metrics) to better supervise targets where DXA under-specifies anatomy. Beyond direct body composition prediction, 3D body scans should be explored for risk-aware metabolic and cardiometabolic applications (e.g., calibrated risk scores, decision support). Studies should also examine longitudinal change detection, quantifying within-person trajectories over time rather than focusing solely on cross-sectional performance. This enables effective patient monitoring and earlier preventive intervention.

## Data Availability

The pretraining datasets are publicly available at the following sites: https://graphics.soe.ucsc.edu/data/BodyModels/ https://humanshape.org/CAESAR/

The remaining datasets contain protected human‐subject information and are available upon reasonable request to qualified researchers who have appropriate approvals and data-use agreements to access confidential data.

https://graphics.soe.ucsc.edu/data/BodyModels/

https://humanshape.org/CAESAR/

## Acknowledgements

The content is solely the responsibility of the authors and does not necessarily represent the official views of the National Institutes of Health.

