## supplemental_file for "BodyMAE: A Surface-Area Aware Masked Autoencoder for Body Composition Estimation from 3D Body Scans"

Cohort characteristics for body composition datasets: age, sex, weight, and height

| | N | Age [mean, std] | Gender | | Weight (kg) [mean, std] | Height (m) [mean, std] |
|---|---|---|---|---|---|---|
| | | | F [size] | M [size] | | |
| NB | 489 | 27.73 (±8.11) | 308 | 181 | 72.47 (±16.91) | 1.71 (±0.09) |
| BA | 252 | 37.71 (±10.09) | 188 | 16 | 114.63 (±23.66) | 1.64 (±0.08) |
| SA | 176 | 76.15 (±6.11) | 128 | 39 | 70.13 (±17.99) | 1.62 (±0.09) |

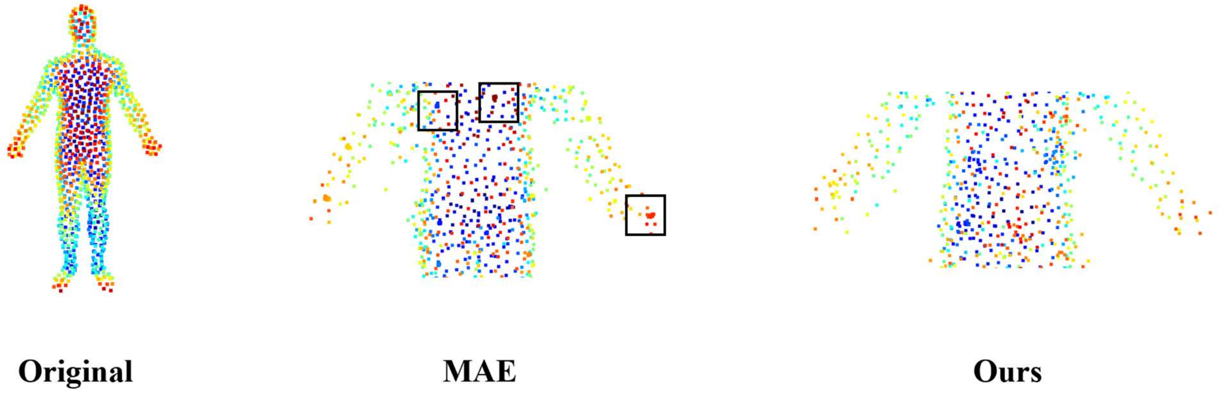


Effect of repulsion loss on reconstructed point distribution. Compared with the baseline MAE, our model enforces repulsion loss to discourage point clustering, yielding a more uniform spatial distribution consistent with the original scan.
